# Joint Prediction of Adjuvant Therapy Response and Time-to-Response for Cancer Patients Using the Personalized-DrugRank Method

**DOI:** 10.64898/2026.03.12.26348235

**Authors:** Fiamma Romagnoli, Marco Pellegrini

**Affiliations:** Istituto di Informatica e Telematica del CNR, Via G. Moruzzi 1, 56100-Pisa (Italy)

## Abstract

**Background:** The ideal of personalized medicine is to support the clinical decision process towards the right drug for the right patient at the right time, by using, among other diagnostic tools, molecular biomarkers that are specifically dependent on the patient status and on the therapeutic options. Several challenges must be overcome to realize this vision. Patients present a wide spectrum of genetic variability even before developing diseases, and disease like cancer add an extra layer of mutations, while only a very small fraction of such variants have diagnostic or prognostic value. Moreover it is also challenging to predict how the patient will respond to a specific drug based on the patient’s ‘omic’ profiling, since any drug introduces further perturbations in the biochemical model.

**Methods:** In this paper we propose the method Personalized-DrugRank for joint prediction of therapy response and time-to-response for cancer patients undergoing pharmacological therapy after surgery. The method is based on personalizing the DrugMerge methodology for drug repositioning in order to extract a few synthetic indices useful as input to ML prediction tools. In particular the proposed methodology is a novel and principled approach to merging independent patient-specific transcriptomic data with drug perturbation data from cell line assays. One of the key novel features of our approach over the state of the art is the joint prediction of the response of the patient to therapy along with an estimate of the time-to-response (i.e the prediction of the time needed for the therapy to succeed or fail).

**Findings:** We tested our methodology on data from the TCGA (The Cancer Genome Atlas) Program for three cancer types (Breast, Stomach and Colorectal cancer), 10 pharmacological regimens and 13 homogeneous cohorts. For the therapy response prediction task we developed models that attain an average AUC performance 0.749, average pvalue 0.030, average accuracy 0.809 with balanced Positive and Negative Predicting Values. For the time-to-event prediction task we developed regression models for the 13 homogeneous cohorts that attain an average (geometric) Concordance Index performance 0.782 (max 0.904, min 0.651) with average log likelihood pvalue 0.004, improving in nine cohorts over 13 upon models based only on clinical parameters having average Concordance Index 0.678 and average p-value 0.006. Interestingly, we attain statistical significant results even with quite small therapy-homogeneous cohorts (ranging from a minimum of 7 patients to a maximum of 32).

**Conclusions:** The ability of predicting with high accuracy the response of a cancer patient to a chosen pharmacological therapeutic regimen along with an estimate of the time-to-response helps adapting the clinical decision process to the specific patient profile, thus increasing the likelihood of providing correct and timely therapeutic decisions.

## 1 Introduction

Cancer is the second leading cause of mortality worldwide and accounts for 277 different types of cancer diseases [Hassanpour and Dehghani, 2017]. According to recent global cancer statistics, the most frequently diagnosed cancer in 2022 was lung cancer (12.4%), followed by cancers of the female breast (11.6%), colorectum (9.6%), prostate (7.3%), and stomach (4.9%) [Bray et al., 2024]. In addition to the variety in site occurrence, phenotypic and functional heterogeneity arise among cancer cells within the same tumor as a consequence of genetic change, environmental differences, and reversible changes in cell properties [Meacham and Morrison, 2013]. This represents a challenge for treatment selection and efficacy due to wide inter-patient variability [Ilan and Spigelman, 2020]. In this context, differential analysis of tumor samples is a key ingredient of a personalized approach. This vision of personalized medicine [Hamburg and Collins, 2010] involves developing and using diagnostic tests based on genetics, proteomics, transcriptomics or other molecular features (often referred to collectively as ‘omics’) to better predict patients’ responses to targeted therapy. Ideally such tests would direct the clinical decision process towards the right drug for the right patient at the right time. The realization of such a vision must overcome, however, many scientific, regulatory and economic hurdles in order to become standard clinical practice. Limiting the discussion to the scientific aspects, it is challenging to identify the molecular biomarkers that have the most relevance to the clinical objectives (either in isolation or in combination with other diagnostic tools, such as imaging). Patients present a wide spectrum of genetic variability even before developing diseases, and disease like cancer add an extra layer of mutations, and thus only a very small fraction of such variants have diagnostic or prognostic value. Moreover it is also challenging to predict how the patient will respond to a specific drug based on the patient’s ‘omic’ profiling, since the drug introduces further perturbations in the biochemical model. Ideally one would like to have drug descriptors obtained from its chemical composition, and from assays on cell lines or organoid tissues obtained before any application of any drug on patients and independently of any specificity of the patients that might take them. Separately, patients with a specific disease should be profiled independently of the possible treatment with the appropriate omics assays and clinical, demographic annotations. Finally the matching of patient and drug profiling should be done completely in-silico. This scenario reduces to a minimum the overhead need and allows to consider both standard and non-standard pharmacological therapies on an equal footing. How to best merge data from such heterogeneous sources is a key technical challenge that needs to be addressed by any algorithm aiming at predicting patients response to drugs [Gligorijević and Pržulj, 2015].

In a seminal paper Iorio et al. [Iorio et al., 2016] using data from multiomic assays of very large collections of cancer samples and cell lines demonstrated that clinically relevant molecular cancer multiomic markers (christened “Cancer Functional Events” (CFEs)) are mostly conserved across patient tissues and cell lines. Moreover, it was shown that such clinically relevant CFE can be used to predict cell-lines drug-sensitivity both singularly and aggregated in combinations through logical circuits, where cell-lines drug-sensitivity is measured by the IC50 dose index of cell viability. These findings underpinned extensive research on cell-line drug sensitivity with the hope that computational models/algorithms performant on such data would carry over to predict therapeutic drug-sensitivity for patients (which can be measured say in terms of overall survival or in terms of therapy outcome). Several computational forecast methodologies have been proposed which are reviewed in [Chen and Zhang, 2021, Xia et al., 2022, Firoozbakht et al., 2022]. While some approaches favor a direct use of raw omics data, others do introduce intermediate layers where one can integrate general biological knowledge from several sources (e.g. from literature digests or from curated pathway databases) in the process. Examples of this second type of approach are [Wang et al., 2019, Kuenzi et al., 2020, Tang and Gottlieb, 2021, Samal et al., 2022, Chawla et al., 2022, Ferraro et al., 2023]. This approach favors interpretability of the predictions and may help when the focus is the identification of new targets for drug development and chemical fine tuning for sub-populations. On the other hand, as pointed out in [Carli et al., 2025] reliance on accrued biological knowledge in the discovery phase may induce biases (e.g those stemming from publication bias). In general lifting results form the cell-line level to the patient level is still an open problem with much ongoing activity (see discussion in Section 3.1).

In this paper we propose the method Personalized-DrugRank (P-DR for short) for joint prediction of therapy response and time-to-response for cancer patients undergoing pharmacological adjuvant therapy. The proposed method strikes a middle ground between the above mentioned approaches to interpretability and prediction performance. We make use of prior biological knowledge in unbiased form as a collection of functional modules from a large gene-coexpression network for *homo sapiens*. Such a modular arrangement has been shown to be effective for uncovering functional modules (validated via GO enrichment as well as with GWAS data) and disease active modules (i.e. those modules that are most affected in a disease defined at a population level) [Lucchetta and Pellegrini, 2020]). Such background is then a framework on which we can integrate drug induced differential expressions from cell lines so to produce repurposable drug rankings [Lucchetta and Pellegrini, 2021]. In the present work we augment further this setting by introducing patient-specific transcriptomic profiling as an additional input to the algorithm, so as to produce *personalized drug sensitivity rankings*. Indices obtained from these rankings are shown to be good predictors of therapy outcomes in terms of AUC, accuracy, PPV and NPV. Indices obtained from these rankings are further validated by using them as input to time-to-event regression models and by measuring their performance with concordance index measures.

Our work can better understood as a progress within the framework of network pharmacology [Nogales et al., 2022] where the objective is to go beyond the one-disease-one-target-one-drug paradigm [Maeda and Khatami, 2018] by simultaneously and implicitly targeting multiple proteins (both on-target and off-target effects) within the space of *a personalized disease modular network*.

## 2 Results

### 2.1 Prediction of therapy response

In the cohorts for which homogeneous treatments result in divergent therapy outcomes we perform a functional analysis as detailed in section 5.7 in order to predict the treatment outcome using the patient’s tumor gene expression profile and the drug perturbation gene expression profile. The method is not using any supervised learning algorithm: it is based on a direct computation of relevant indices and on a threshold to assign the patients to high risk (prediction of Progressive Disease/Stable Disease) and low risk (prediction of Complete Remission/Partial Remission) classes. In Table 1 we report the AUC and AUC pvalue as well as the maximal accuracy attained by an optimal threshold with associated PPV and NVP values. Statistical significance is attained for AUC in all but one cases, notwithstanding the small number of patient in each cohorts. Accuracy ranges for a maximum of 0.95 and a minimum of 0.73, with average 0.80, and with well balanced NPV and PPV in all experiments. The Reactome database of functional annotation attains better results for AUC and maximal accuracy in two cases out of three. In practice both functional annotated database should be assessed to select the preferred one in a specific cohort. For the CRC-Irinotecan cohort we attain comparable AUC and mACC results using as query drugs Fluorouracuil and Irinotecan; this observation indicated high consistency of the methodology since both drugs are administered in this cohort. Overall for each cohort, we attain significant results for at least one of the two biological process annotation databases, with high accuracy (about or above 0.8) and good balance between NPV and PPV values. Given the small numbers involved in each cohort, attaining statistical significance is challenging and these results indicate that the Personalized-DrugRank algorithm is effective in uncovering the relevant features of the prediction problem in the mechanistic model.

**Table 1:**
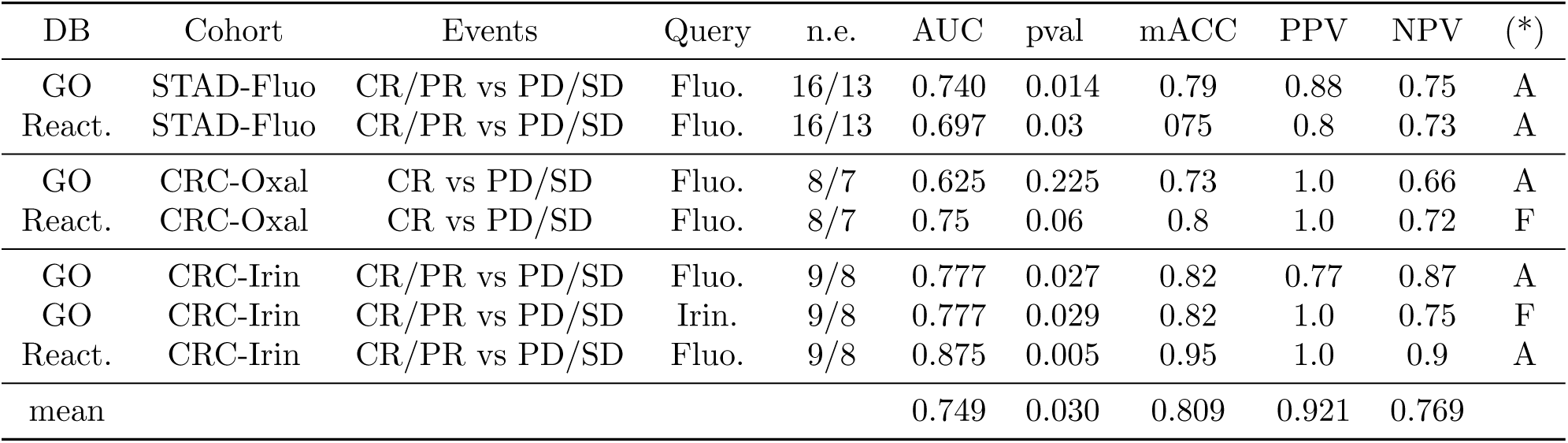
Event Prediction for homogeneous cohorts by therapy. GO stands for Gene Ontology Biological Process database, React. stands for the Reactome Database. The cohort STAD-Fluo is formed of Stomach Adenocarcinoma patients receiving Fluorouracil only as therapy. CRC-Oxal is formed of Colorectal Adenocarcinoma patients receiving Fluorouracil and Oxaliplatin as therapy. CRC-Irin is formed of Colorectal Adenocarcinoma patients receiving Fluorouracil and Irinotecan as therapy. Query indicates the drug for which the prediction parameters are reported (corresponding to one of therapy drugs given to the patients). mACC is the maximal accuracy for an optimal discriminating threshold. Note (*): Method of p-value filtering: (A) Adaptive, (F) Fixed. For p-values the geometric mean is used.

### 2.2 Prediction of time-to-response

Tables 2 (BRCA) and 3 (STAD+CRC) give an overview of the performance of Cox regression models predicting the time-to-event when fed with Personalized-DrugRank indices as part of their input along with standard clinical parameters. Overall the experiments cover three cancer types (Breast Cancer Adenocarcinoma (BRCA), stomach cancer adenocarcinoma (STAD) and Colorectal Cancer (CRC)) over ten therapeutic regimens, resulting in 13 different homogeneous cohorts (both in treatment and outcome).

As the main regression method we use Cox Proportional Hazard model (parametric and semi-parametric). We optimize on 42 different configurations of clinical-only features and on 128 configurations of mixed clinical and P-DR configurations. In Table 2 we report the p-value of the Log-likelihood test for the best model and the best k-fold validation mean Concordance Index (for different values of k) and standard deviation over a number (10) of repeated randomized data-splits.

**Table 2:** Summary of Cox regression survival analysis results for BRCA treatment sub-populations. The event is Complete Remission/Partial Remission in all cases. [Subpop. A] Includes patients receiving Paclitaxel and Cyclophosphamide but not Fluorouracil or Docetaxel. [Subpop. B] Includes patients receiving Cyclophosphamide but not Fluorouracil. [Subpop. C] Includes patients receiving Cyclophosphamide and Fluorouracil. [Subpop. D] Includes patients receiving Cyclophosphamide and Docetaxel. [Subpop. E] Includes patients receiving Cyclophosphamide, Paclitaxel, and Doxorubicin Hydrochloride. pval is relative to the log likelihood test. CI is the mean concordance index in a k-fold validation test over 10 randomized repetitions of the k-fold validations. ST indicates subtype present in the best model. For p-values and CI the geometric mean is used.

| Group data |  |  |  | clinical+PDR |  |  |  | clinical only |  |  |  |
| --- | --- | --- | --- | --- | --- | --- | --- | --- | --- | --- | --- |
| Subpop. | key | n.o. | n.e. | CI | std | pval | N. | CI | std | pval | N. |
| A | Pa. | 27 | 23 | **0.880** | 0.081 | 0.0007 |  | 0.708 | 0.176 | 0.18 | ST |
| B | Cy. | 35 | 32 | 0.736 | 0.318 | 0.017 |  | **0.777** | 0.152 | 0.02 |  |
| C | Fl. | 12 | 11 | **0.854** | 0.147 | 0.01 |  | 0.761 | 0.132 | 0.32 |  |
| D | Doc. | 22 | 22 | 0.708 | 0.248 | 0.37 | ST | **0.735** | 0.08 | 4.9E-7 |  |
| E | Dox. | 17 | 16 | **0.904** | 0.143 | 0.002 |  | 0.806 | 0.300 | 0.006 |  |
| mean |  |  |  | 0.812 |  | 0.009 |  | 0.756 |  | 0.005 |  |

Critical to attaining precise predictions is the subdivision the BRCA cohort into subpopulations receiving a homogeneous pharmacological treatment. For the 5 homogeneous sub-population by treatment we attain almost always significant p-values and high concordance values. For the STAD and CRC patient cohorts we perform similar experiments, where we also explore homogeneous sub-populations by uniform treatments when it is suitable (Table 3). Both in Table 2 and in Table 3 we compare the Concordance Index found with a mix on P-DR indices and clinical indices with the best CI obtained using only clinical parameters and we found an increase in performance In 9 cases, worse performance in 3 cases, and a tie in one case. Overall these measurement demonstrate an independent predictive power of the P-DR indices w.r.t the standard clinical parameters in a significant number of scenarios. The log-likelihood test p-value is most of the times significant, which is remarkable given the relatively small number of patients of the homogeneous cohorts.

**Table 3:** Summary of Cox regression survival analysis results for STAD and CRC treatments. pval is relative to the log likelihood test. PD is Progressive Disease, CR is Complete Remission, PR is Partial Remission, and SD is Stable Disease. pval is relative to the log likelihood test. CI is the mean concordance index in a k-fold cross-validation test over 10 randomized repetitions of k-fold validation (default). ST indicates subtype present in the best model. i20 indicates that 20 random repetitions of k-fold cross-validation has been used. For p-values and CI the geometric mean is used.

| Group data |  |  |  |  | clinical+PDR |  |  |  | clinical only |  |  |  |
| --- | --- | --- | --- | --- | --- | --- | --- | --- | --- | --- | --- | --- |
| Cohort | Event | key | n.o. | n.e. | CI | std | pval | N. | CI | std | pval | N. |
| STAD | CR | Fl. | 28 | 15 | **0.776** | 0.208 | 0.0005 | ST | 0.713 | 0.282 | 0.01 |  |
| STAD | PD | Fl. | 28 | 12 | **0.819** | 0.201 | 0.017 | ST | 0.666 | 0.08 | 0.008 | ST |
| CRC | CR/PR | Fl. | 9 | 9 | **0.813** | 0.266 | 0.03 | i20 | 0.410 | 0.110 | 0.06 | i20 |
| CRC | CR | Ox. | 15 | 8 | **0.788** | 0.173 | 0.023 | i20 | 0.591 | 0.08 | 0.49 | i20 |
| CRC | PD/SD | Ox. | 15 | 7 | **0.793** | 0.143 | 0.001 | i20 | 0.765 | 0.303 | 0.08 | i20 |
| CRC | CR | Le. | 17 | 16 | **0.651** | 0.276 | 2.1E-7 |  | 0.514 | 0.12 | 1.7E-8 |  |
| CRC | CR/PR | Ir. | 17 | 9 | 0.834 | 0.225 | 0.02 | i20 | 0.833 | 0.082 | 0.01 | i20 |
| CRC | PD/SD | Ir | 17 | 8 | 0.665 | 0.230 | 0.15 | i20 | **0.692** | 0.154 | 0.42 | i20 |
| mean |  |  |  |  | 0.764 |  | 0.002 |  | 0.633 |  | 0.007 |  |

### 2.3 Role of cancer subtypes

Often molecular subtypes of cancer are used as a qualitative indication useful for therapy selection and prediction of therapy outcome and timing [Karam et al., 2025]. In our cohorts for colorectal cancer (CRC) only one subtype is present thus subtyping is not a useful discriminatory variable and the role of molecular subtypes could not be measured for CRC. For the breast cancer cohorts (BRCA), results in Table 2 indicate that molecular subtype features are not critical to attain the best performance. In contrast Table 3 indicates that for Stomach Cancer (STAD) molecular subtype features improves the predictive power of regression models. Thus the question whether it is an advantage the use of subtyping in time-to-event modeling should be examined in dependence of the single disease/cohort under consideration.

## 3 Discussion

### 3.1 State of the art

Personalized medicine [Hamburg and Collins, 2010] has been one of the leading themes of recent research in biomedicine, with the aim of matching extensive knowledge of genomic human variability with increasing accrued information on drugs and diseases so as to improve the quality of evidence-based clinical decisions. Costello et al. [Costello et al., 2014] report on the results of a DREAM challenge (NCI-DREAM7 challenge - 2012-2014) that aimed at predicting the IG50 dosage (the dosage that inhibits cell growth by 50% after 72 hours of treatment) in 53 breast cancer cell lines (35 for training, 18 for testing) across 38 drugs. The main conclusion of this study is that for most drugs and most cell lines predictive signals can be identified within omics measurements (including RNAseq, Gene expression by microarray, Methylation, CNV and RPPA assays). This study, along with Iorio et al. [Iorio et al., 2016], gave impetus to the idea of transferring cell line drug sensitivity prediction methods to the analysis of more complex patient data.

Due to the importance of the topic several surveys and literature reviews have appeared in recent years (see e.g. [Adam et al., 2020], [Feng et al., 2021], [Panja et al., 2021], [Firoozbakht et al., 2022], [Kong et al., 2022], [Shen et al., 2023]). Evaluating personalized predictions of drug response in a clinical setting is a challenging task in its own as discussed in [Ding et al., 2016] and [Scarpato et al., 2024] due to paucity/sparsity of data and to the retrospective nature of the data collected. He et al [He et al., 2018] resort to an indirect validation methodology based on matching the patients’ breast cancer hormonal profile with the class (hormonal, chemotherapic) of the predicted effective drug.

The recent rise of Deep Neuronal Networks learning techniques has produced a new flurry of research in this area of personalized recommendations [Partin et al., 2023], [Katzman et al., 2018], [Liu et al., 2022], [Wang et al., 2021] and [Jia et al., 2021]. Here we skim through relevant recent results, highlighting the proposed algorithmic approach to the problem of merging cell line data with patient data, and the type of data used as input to the prediction tool.

Carli et al. [Carli et al., 2025] use large-scale pharmacogenomics databases, including PRISM and GDSC databases, along with the output of systematic LLM interrogations on putative drug mechanism of action (MOA) to train a pool of ML models on the task of predicting IC50 values for cell lines as target variables. Such trained models are then modulated, via the *Celligner* [Warren et al., 2021] tool, to take TCGA pancancer patient data as input. The analysis encompassed 9805 TCGA samples and 41 clinically relevant drugs, with the aim of finding whether for any drug-tumor type pair the relative number of patients receiving the drug is statistically correlated with a high ranking in the IC50 target prediction ranking. Note that the actual response of patients to the drugs is not the focus of their analysis.

Su et al. [Su et al., 2022] proposed a deep learning-based method, named Siamese Response Deep Factorization Machines (SRDFM) Network, in which both drug property and gene expression were adopted to build a concatenated feature vector, this enables the recommendation for newly designed drugs with only chemical property known.

Among the methods using only clinical data from Electronic Health Records (EHR), Bohi et al. [Bhoi et al., 2021] use large databases of clinical and demographic data from EHR (MIMIC-III is a very large publicly available EHR dataset, which contains clinical data for 7,870 neonates (infants) and 38,597 adults admitted to the ICU between 2001 and 2008) in order to provide indication/suggestions for emergency treatments on new ICU patients.

Chawla at al. [Chawla et al., 2022] introduce *Precily*, a Deep Neural Network (DNN) trained to infer treatment response in cancer using in training cell-lines gene expression data (both bulk and single cell RNAseq), moreover, *Precily* models pathway activity estimates in tandem with chemical drug descriptors as input features. The method is shown to be an effective predictor of cell-line drug response (IC50 values) as well as of drug response in cell line–derived xenograft (CDX) models for prostate cancer receiving treatments to induce castration-resistance (CRPC) followed by treatment with enzalutamide (ENZ). *Precily* has been applied then to a multi-cancer pool of TCGA patients (1443 unique patients over 29 cancer types, with 139 unique drugs, and 3108 patient-drug combinations with recorded clinical responses) with 90% of the data used for training and parameter tuning and 10% for testing, resulting in AUC-PR of 0.77. The same data fitted with an algorithm ‘Extremely Randomized Trees’ (XRT) from the AutoML suite produced a final performance evaluation of AUC-PR of 0.85.

Peng et al. [Peng et al., 2022] deal with the problem of the merging of cell-line and patient data at the level of the gene-expression matrix by considering it a problem akin to that of batch-correction. Sinha et al. [Sinha et al., 2022, Sinha et al., 2024] have shown that useful predictions in precision medicine can be obtained by adapting bulk-rnaseq cell lines models to work on sc-rnaseq data and patient-derived cancer primary cells (PDC) clone drug response assays.

Ma et al. [Ma et al., 2021] propose to treat the problem of transferring predictive power from cell-line trained models to patient data as a problem in machine learning termed *few-shots learning* that is way to realize *transfer learning*: an approach that postulates that prior knowledge acquired in one problem domain can be reused and applied to solve different but related problems. Adapting the input numerical values from heterogeneous domains is attained by common reference to the normal distribution via z-scores. The resulting algorithm *translation of cellular response prediction (TCRP)* is then tested on increasingly difficult benchmarks involving cell line data, Patient Derived Tumor cells cultures in vitro (PDTCs), and PDX-derived cell lines. The main emphasis in the analysis is to determine how many shots are needed in order for the learning process to reach stability. Experiments on PDX-derived cell lines (the situation closer to the aim of response prediction for patient) were conducted on data for 228 PDX mouse models in the GDSC1000 dataset, where each model was exposed to one of the five drugs on which TCRP had been trained in cell lines (cetuximab, erlotinib, paclitaxel, tamoxifen and trametinib). After dichotomizing predictions and response to a common matching scale, TCRP attained the following accuracy in response prediction: cetuximab (0.63), paclitaxel (0.64), tamoxifen (0.73), trametinib, (0.70), erlotinib (0.71). A follow up study by So et al. [So et al., 2023] applied TCRP to two large clinical trial patient datasets for breast cancer (GSE25066 receiving neoadjuvant taxane-anthracycline chemotherapy and GSE41998 receiving neoadjuvant Doxorubicin/Cyclophosphamide followed by Ixabepilone or Paclitaxel, with resp. 508 and 279 samples). TCRP remains competitive w.r.t four baseline standard ML method (Nearest Neighbor, Neural Network, Random Forest, and Linear Regression), however global predictive performance measured by the correlation coefficient drops from about 0.3 as measured in [Ma et al., 2021] to 0.25 and 0.1 in the two patient cohorts. Venezian Povoa et al. [Venezian Povoa et al., 2021] propose the Multi Learning Training approach (MuLT) method combining supervised, unsupervised and self-supervised learning algorithms to predict heterogeneous treatment outcomes for Multiple Myeloma (MM). This scheme uses as input gene expression values as well as genetic abnormalities detected by Fluorescence in situ hybridization (FISH) testing. The ROC AUC performance attained ranges from about 0.56 to 0.69 for the several models proposed.

Sotudian et al. [Sotudian and Paschalidis, 2021] use cell line data and drug sensitivity data from the Cancer Cell Line Encyclopedia (CCLE) [21] and the Cancer Therapeutics Response Portal (CTRP v2) to predict relative response of a drug-cell line pair against an alternative drug-cell line pair. Concordance index performance on three data sets (Myeloma+lymphoma, blood, and lung) range from 0.73 to 0.64 for the proposed method over a set of 50 drugs. This article does not discuss transferring the model trained on cell lines data towards applications for personalized predictions using single cancer patients data.

Gerdes et al. [Gerdes et al., 2021] present an approach, named Drug Ranking Using ML (DRUML), which uses omics data to produce ordered lists of more than 400 drugs based on their anti-proliferative efficacy in cancer cells. DRUML is trained on proteomics and phosphoproteomics data derived from 48 cell lines, and it is verified with data comprised of 53 cellular models using as target for prediction the Area Above the drug dose–response Curve (AUC) showing the fraction of surviving cells as drug concentration increases (with the median AUC cutoff value for dichotomization). For applicability to a clinical setting they show that a pool of AML patients predicted to be sensitive to cytarabine by DRUML shows greater clinical overall survival (OS) than those predicted to be resistant (Median OS was 1.1 vs 3.4 years (p = 0.0049, n = 10 and 15 respectively).

Mourragui et al [Mourragui et al., 2021] solve the problem of merging patient and cell line data with the TRANSACT algorithm, a computational framework that uses both standard linear PCA and non-linear (Kernel based) principal components analysis to map both cell-line drug perturbation gene expression and patient tumor samples gene expression in the same high dimensional geometric space where dimensionality reduction and low-rank interpolations are performed and optimized to reduce the total remapping error. The final result allows us to integrate via this mapping the two data sets into a unique predictor schema. Kuenzi et al. [Kuenzi et al., 2020] develope DrugCell, an interpretable deep learning model of human cancer cells trained on the responses of 1,235 tumor cell lines to 684 drugs. The Drugcell method is based on feeding a NN model with drug chemoinformatic representations (specifically the Morgan Fingerprints, a 81-bits 0/1 vector) ad a cell status represented as a 0/1-genotypic mutations vector for the top 15% most mutable human genes. Interestingly this genomic profiling is sufficiently at a high level so to be usable as is also to represent the mutation status of single patients’ cancer tissues. The derived pretrained model is then tested on a large collection of Patient Derived Xenograft(mouse) models, where DrugCell classifies pairs of PDX-drug as respondent or non respondent. This classification is then evaluated and a statistical significant (log likelihood test) association is found with a better average PFS for the respondent class. Finally the pre-trained model is also applied (as is) to a cohort of 221 estrogen receptor (ER)-positive metastatic breast cancer patients who had undergone multiple rounds of therapy, including an ER antagonist (fulvestrant) in addition to treatment with an mTOR inhibitor (everolimus) or CDK4/6 inhibitor (ribociclib). Here too DrugCell classifies the patients into respondent and non-respondent classes to the specific drugs administered to the patients. For this stratification, there is a statistical significant (log likelihood test) association with better PFS for the respondent class. Naturally, performing this type of stratification does automatically imply the ability to do point-wise predictions for any single patient with a reasonable accuracy.

Kim et al. [Kim et al., 2019] consider the translation problem as a problem in dimensionality reduction at the level of genomic features. The propose a method is based on parallel factor analysis with additional constraints so to identify the factors (genomic feature) most important for drug-response prediction in multiomic cell line perturbation data, which at the same time perform well on patient-derived xenografts (PDX) data.

Huang et al. [Huang et al., 2018, Huang et al., 2017] use a ML schema based on SVM to predict dichotomized drug responsiveness of patients using data from the TCGA databases. Predictors for two drugs (gemcitabine (92 patients) and fluorouracil (60 patients)) are built using patients affected by a variety of cancer types. Evaluation using a LOOCV schema gives the following results: ACC GEM 81.5%, PPV GEM 77.8%, NPV GEM 83.9%, ACC 5-FU 81.7%, PPV 5-FU 83.3%, NPV 5-FU 79.2%. Note that Huang et al. use only patient cancer samples RNAseq data and do not make use of data from cellular lines, nor of the patient standard clinical parameters. No prediction of time-to-event is reported in [Huang et al., 2018, Huang et al., 2017].

Our review of the state of the art focuses on methods that are based on molecular and clinical data. Other approaches are possible, and in particular radiology-based imaging can be used to monitor and predict targeted therapy response in cancer [Yang et al., 2025a]. Advantages of radiology-based imaging are its low impact and the possibility of repeated measurements during the treatment. On the other hand the models proposed in such studies do not typically make use of drug-profiling data and require ad-hoc algorithmic re-tooling and training for each cancer type and therapeutic regimen.

In summary, the overview of the state of the art confirms that our methodology for merging cell line and patient data in a common model is novel and it is able to bridge well the methodological gap between performing prediction on cell lines and other patient-derived models, versus the actual patient response. Moreover while most existing methods aim at just a categorical prediction (or a population stratification), our method extends to accurate time-to-event predictions.

### 3.2 Open problems

As drug response prediction is a vast subject, with implications from several areas of biology and medicine, we comment on the relationship of Personalized-DrugRank with several issues arising in the literature. Each issue is introduced by a heading.

#### Mechanistic models vs. trained models

Most work for drug response prediction is done in a classical supervised ML setting (either using deep NN methodologies or other ML methods). This gives emphasis on raw predictive performance at the expense of providing clues to the underlying biological processes governing the drug effectiveness (i.e. a plausible candidate MOA to be eventually tested in ad-hoc assays) which is important for drug refinement phases [Gherman et al., 2023], [Baker et al., 2018]. Our approach is intermediate, in the first phase the P-DR algorithm works in a purely direct manner (thus closer to a mechanistic view) in order to merge DEG lists from cell-line drug effect data and patient diseased tissues data within the context of a large co-expression network to extract a few personalized drug effect indices. In the second phase such indices are used in conjunction with classical clinical indices in order to produce regression models for the prediction of drug-induced outcome and time-to-event.

#### Bulk vs Single Cell data

Transcriptomic data is mostly collected on tumor samples via bulk RNAseq or microarray technologies. These assays are able to produce an average landscape of the tumor’s gene expression at a specific point in time. It is however well known that tumors are often composed of several clones, each with a more specific profiling and often the insurgence of drug resistance is due to the proliferation of under-represented clonal cell lines over the treatment time period. The emergence of novel single cell omic profiling technologies for the identification of tumor clones may be a way to better characterize such variability (see e.g. the work of Sinha et al. [Sinha et al., 2022, Sinha et al., 2024] and Wang et al. [Wang et al., 2022]. The potential gains due to such deeper assays must be weighted against the more complex and more expensive procedures involved. In particular in this paper we show that bulk RNAseq data can provide results (in terms of accuracy, PPV/NPV and AUC values) at a par with the best results in [Sinha et al., 2022] attained with SC data.

#### Single cell transcriptomics, clones and resistance

Our experiments are based on patient’s tumor sample transcriptomic data obtained via bulk mRNA-seq or microarray assays. It is well known that such gauging methods average expression readings across the tumor’s clonal composition. On the other hand, clonal cell sub-population with a minority representation at the time of sample collection may be responsible for the emergence of drug resistance (according to one of the accepted explanations for the emergence of drug resistance). In this setting our methodology might miss the signal representing the potential drug resistance future phenomenon. To overcome this difficulty we speculate that if sc-RNAseq data from a patient tumor sample is available at the clonal sub-populations level, we may replicate our analysis on each clonal cell subpopulation separately and thus differentiate the clonal sub-populations most likely to respond to a candidate drug versus those less responsive [Liu et al., 2025]. For these less-responsive sub-populations the system might suggest alternative drugs (among those for which cell-line response data is available) more likely to be effective for that specific clonal cell sub-population, to be used in a multi-pharmacological therapeutic strategy.

#### Disregarding the MOA

Our approach concerning drugs does not make use of explicitly known drug MOA (this is in contrast with say [Carli et al., 2025, Ma et al., 2021] where knowing the main protein/gene/pathway targeted by a drug is instrumental in the initial gene feature selection phase). Instead, we quantify global perturbation effects of a drug on the whole patient-specific disease active sub-network (with its subdivision into active modules). This choice has several advantages. First of all the MOA might be unknown, it might be poorly defined, uncertain or disputed. Even when a MOA is known (and even when a drug is designed for a specific MOA or molecular target) it is still possible that the reason of effectiveness of the drug is only weakly associated with it [Lin et al., 2017, Lin et al., 2019]. In particular often many off-target effects might be as relevant, but less understood or even identified. Our implicit approach does not rely on precise characterization of MOA and thus potentially captures both on-target and off-target effects.

#### Extension to non-cytotoxic therapies

In the context of oncology the main effect of drugs is often to induce cell death in cancerous cells while preserving normal cells from the same fate. In this context it is coherent to take IC50 cell viability measurements in cancer cell lines as a potential relevant measure of the drug’s effectiveness in patients. However reliance on IC50 cell viability measurements is less relevant for other scenarios. For example in metabolic disease we might try to use drugs to re-balance disrupted molecular pathways without resulting in cell death. Our approach (that does not rely in cell viability measures) is still in principle justified also in this non-oncological scenario.

#### Explainability and MOA

While we do not rely on knowing a MOA in order to perform time-to-event prediction we do recognize that uncovering the MOA that are effective at the level of single patients is an important step towards refining the therapeutic choices, and maybe finding more effective drug combinations. In this scenario our method can readily provide the relevant functional annotations.

We support drug MOA discovery by checking for each patient and each drug the personalized disease modules most contributing to the final score (sorted in order of importance). Such modules can then be analyzed for their enrichment in known pathways (e.g. KEGG or GO annotations) thus providing a weighted list of plausible MOA for the drug in the specific patient. Note that in this framework it is easy to uncover synergies among multiple drugs towards a common effect, for example when different drugs affect alternative molecular pathways for the same function.

#### Drug toxicity

Most chemotherapeutic agents are cytotoxic to rapidly dividing cells, thus toxicity primarily affects all body tissues with high cell turnover. The proposed Personalized-DrugRank methodology takes as input measurements form cultured cell lines (for drug perturbations) and from the patient primary cancer tissue samples (for personal cancer profiling) thus our method does not capture explicitly toxicity effects occurring in the patient when computing the drug-related indices. We speculate that toxicity can be more readily included in our predictive models by incorporating in a Cox regression model specific genomic biomarkers (correlated with the drug susceptibility in multiple body tissues) and/or global health parameters.

#### Alternative end-points

In this work we use as end-point the therapy response (more specifically to the first line of treatment) and we also predict the time-to-event (often also termed time-to-failure (TTF) when the event of interest is the failure of the chosen therapy). Initial test of Personalized-DrugRank with alternative end-points, such as progression free survival (PFS) and overall survival (OS), did not produce satisfactory results. A possible explanation for this observation is that typically cell-line transcriptomic assays found in public databases (GEO) measure the drug action on cell lines in a relatively short time frames (rarely above 72 h), thus they might miss measuring some longer-term effects that are relevant for the phenomenon of tumor relapse (local, regional or distant) occurring after therapy has attained the desired effect of cancer remission. An additional effect that we are not considering at the moment, but that influences heavily both PFS and OS, is the status of the tumor micro-environment that is typically not a part of the patient sample omic analysis [Almazrouei et al., 2025].

#### On the number of needed molecular biomarkers

Transcriptomic gene expression data in the TCGA (The Cancer Genome Atlas) Project is generated via RNA-Seq technology ( Illumina high-throughput sequencers) that measure simultaneously about 20K protein coding genes (Level 3 / GDC harmonized data) [Weinstein et al., 2013]. A similar order of magnitude in the number of protein-coding gene expression levels is available from the GEO (Gene Expression Omnibus) Drug perturbation Database (via a variety of technologies like microarray and RNA-seq assays). Since the cost of collecting data for the drug perturbations on cell lines is incurred only once per drug/cell line pair, the cost of this data collection can be amortized across all the patients for which the predictions are computed. On the other hand, the cost of collecting the patient-specific cancer gene expression data is incurred on each single patient thus it is important to attentively consider the cost associated with this part of the data collection. One possible approach to cost reduction could be the application of High-throughput landmark gene hybridization with inferred expansion (L1000 Assay), which involves experimental measure of roughly 1000 landmark genes, and computational inference of the expression level of the remainder protein coding genes [Subramanian et al., 2017]. This approach strikes a balance between cost reduction and general applicability of the methodology to a wide range of cancer types. The problem of further reducing the number of measured molecular biomarkers is left as an open problem for future research. It is likely that the relevant biomarkers will be different in each cancer type and thus larger cohorts of patients will be needed in order to determine few, reliable, statistically significant cancer-specific molecular biomarkers that lead to approximating the proposed P-DR indices. Such few biomarkers will be readily measurable via low-cost technologies such as Real Time PCR, available in most clinical settings [Wang et al., 2026].

### 3.3 Relevance in clinical practice and clinical trials

#### Possible Uses in Clinical Trials

One persistent problem in designing clinical trials meant to compare and measure the statistical difference in outcome between a standard treatment (A) against a novel proposed treatment (B) is that of selecting the patients eligible for the trials, and in this pool of patients decide which patients should undergo treatment (A) and which should receive treatment (B). While one can certainly decide such protocols *a priori*, this way of approaching the problem is in contrast with an ethical duty of providing the best available and evidence-based cure to each patient individually. Thus it is gaining popularity a drive towards adaptive trial designs and real-time monitoring leading to adjusting trial parameters based on interim results [Fang et al., 2025]. A prediction system like Personalized-DrugRank, is able to produce statistically significant predictions (for both the event and the time-to-event) with a few dozed cases (in contrast with most of the supervised ML approaches that attain good performance only after training with hundreds of cases). Having the time-to-event prediction helps in switching in timely manner the therapy for the single patient if the predicted beneficial effect of a therapy does not materialize within the predicted time-frame (thus indicating timely a possible failure of the prediction). Interestingly, even if the event prediction is Complete Remission for both treatments (A) and (B), the clinician can use the time-to-event as a further important decision parameter, giving priority to the treatment that is predicted to take the least time to produce its effect, with a likely better trade-off between dosage and toxicity effects.

#### Possible use of Personalized-DrugRank in clinical practice

While there are many methods and algorithms in the literature that tackle the task of predicting patient response to drugs, with a wide spectrum of associated performances, to the best of our knowledge this is one of few methods that, besides event prediction, tackles a joint prediction of the time-to-event for the chosen drug-based therapy. In particular it is possible to take advantage of the fact that Personalized-DrugMerge requires fewer cases for producing statistically significant models, thus it can be applied in principle to small cohorts of patients receiving homogeneous treatments. In order to analyze existing therapeutic protocols we can use the patients accrued clinical records, along with proper transcriptomic analysis of stored tissues if these have been stored for long term conservation.

Such more refined information/prediction may influence the ongoing personalized therapeutic choices for new patients in several ways. Besides providing support for the choice of a drug (or drug combination), it helps in setting up a monitoring time-schedule with the aim of early detection of deviations from the predicted time-to-event. In case of large deviation or insufficient progress, the clinicians might decide an early switch to an alternative drug regimen in a more timely manner, and thus with likely increase in survival chances for the patient [Zhou et al., 2020, Jiang et al., 2024, Yang et al., 2025b].

## 4 Conclusions

In this work we have proposed a principled way to merge drug-related transcriptomic data from cell lines with patient-specific transcriptomic data and thus produce a few synthetic indices useful in predicting the individual response of a patient to a drug (via simple thresholding decision) and the time-to-response (via Cox Regression models that integrate also clinical and subtyping data). The main finding is that these indices appear to be independent of other standard clinical parameters, thus lead to an improvement of the state-of-the-art in personalized predictive medicine. Moreover the indices are obtained in a completely mechanistic prospective which leads to a more robust generalizability of the techniques to other cancer types and other diseases. Both thresholding decision and Cox Regression models are easy-to-interpret models which helps overcoming the possible clinician skepticism owing to the challenges with interpretability of ‘black box’ recommendations. Future research directions are the study of models including routine cancer-specific parameters (e.g Gleason scores in Prostate Cancer, or Homologous Recombination Deficiency (HRD) in Ovarian Cancer) interacting with the novel Personalized-DrugMerge indices. The question whether the prediction power of Personalized-DrugMerge can be preserved when only a few molecular biomarkers are measured using RT-PCR (thus avoiding more expensive and complex assays) is also worth pursuing.

## 5 Methods

### 5.1 Method overview for prediction of time-to-response

The present work builds upon two previous publications [Lucchetta and Pellegrini, 2020] and [Lucchetta and Pellegrini, 2021]. In [Lucchetta and Pellegrini, 2020] it is shown how to adapt the graph community detection algorithm Core&Peel [Pellegrini et al., 2016] to the task of finding functional modules and disease-specific active subnetworks in a large gene co-expression network. The application was validated against the methods participating in the Disease Module Identification DREAM challenge [Choobdar et al., 2019], as well as other state-of-the-art algorithms for disease-specific active subnetwork detection. In [Lucchetta and Pellegrini, 2021] the modular disease-specific active subnetworks of [Lucchetta and Pellegrini, 2020] are combined with data from drug-induced differential gene expression repositories (mainly form assays on cell lines) into a scoring/ranking scheme so to produce ranked list of drugs likely to be effective for the target disease. The proposed algorithm DrugMerge was evaluated versus competing ranking methods in its ability to detect drugs in clinical use high in the ranking. The rationale of that study was that of using novel drugs high in the ranking as candidates for drug repurposing for the specific disease, and at the same time use high ranking drugs already in use as evidence of the global rank’s quality.

In the present research we change the focus and we show that a modification of DrugMerge (Personalized-DrugRank) can be used to make predictions on the efficacy of a specific drug (or combination of drugs) for individual patients affected by the target disease. Thus we shift from making generic predictions of efficacy for a drug in a specific disease to making prediction of efficacy of a drug for a specific patient affected by the disease.

The methodology has two phases. In the first phase Personalized-DrugRank is used to produce four patient-specific numerical indices for each patient that capture the effect upon the personal disease-specific active subnetwork of the drugs used generically for the disease and the drugs used effectively or to be used on the single patient.

The second phase uses classical survival regression models (Cox Proportional Hazard) to incorporate both standard clinical indices (Lymph node stage, Tumor stage, Metastasis stage, and Disease Grade) and the computed P-DR indices into a common regression model for the time-to-event prediction. A schematic figure of the Personalized-Drugrank pipeline for *time to response* calculations is in Figure 1.

**Figure 1:**
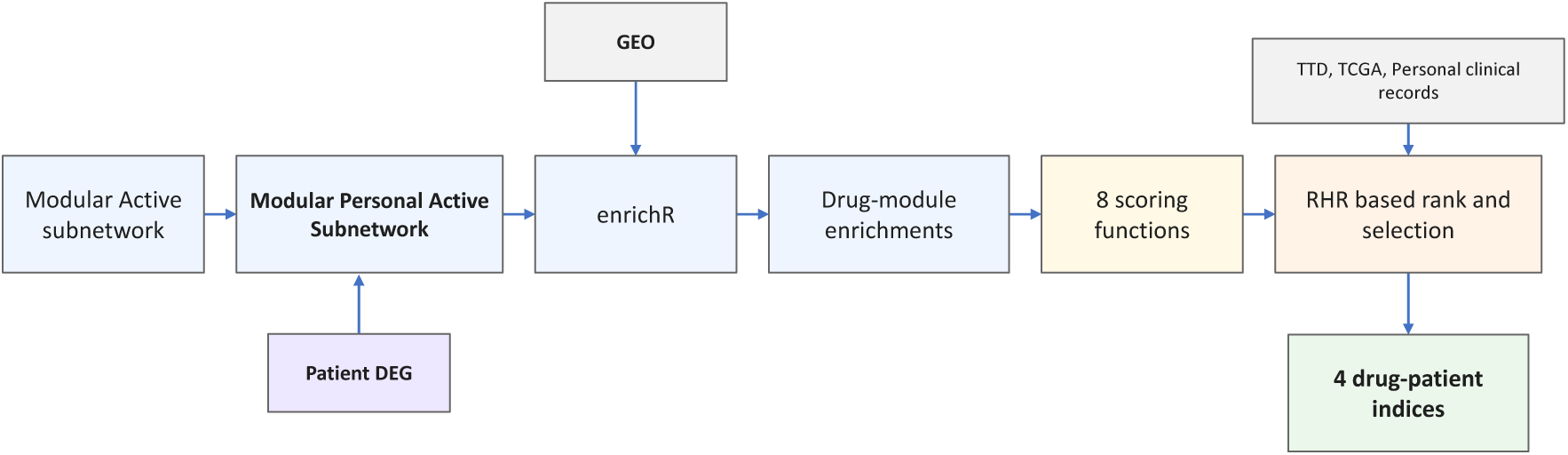
Pipeline for Personalized-DrugRank Index Computations for Time-to-response prediction.

### 5.2 Personalized Indices

Personalized-Drugrank differs from DrugMerge in two key points: 1) The input to Personalized-DrugRank is a list of DEG where we compare genes expressed in the patient’s cancer tissue with the gene expression in a pool of sane tissues from a sane cohort. This input captures the patient-specific DEG. 2) While DrugMerge uses only two scores to rank the drugs against the disease active modules, we use a pool of 8 scores to produce 8 rankings. After evaluating the quality of each ranking against the pool of disease-specific drugs by RHR (Reciprocal Hit rank) we select the ranking having the highest value of RHR for computing the drug-patient indices.

The Reciprocal Hit-Rank (RHR) [Deshpande and Karypis, 2004] is defined as follows. For a list *L* of predictions length *N*, let the hits be in positions *p*_1_, *p*_2_, …,*p_h_*. We define :

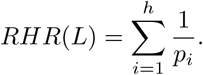

Note that this formula does not imply a fixed length for the lists being compared, and it is fair in comparing both long and short lists. A longer list may have more hits by chance, however it will have positive but diminishing returns for hits in its tail.

The four indices are as follows:

- *RHR score* is the value of RHR used in selecting the ranking of interest (highest among the RHR values)
- *Global rank* is the ranking position in the selected ranking of the highest drug among those used generically for the disease, normalized on the number of such drugs in the reference database.
- *Local rank* is the ranking position in the selected ranking of the highest drug among those used specifically for the patient, normalized on the number of such drugs in the reference data base.
- *DrugMerge prediction* is a binarized version of Global rank where we give value 0 if the normalized Global rank is below 10%, 1 otherwise.

### 5.3 Patients DEG filtering

As patient DEG analysis tends to produce a large number of differentially expressed genes even after filtering for FDR with threshold at 0.01, we introduce further filtering by separating the up regulated and the down regulated genes. Separately in each list we sort the DEG by the absolute value of the fold change and we take the sublist comprising the top 4000 genes in each list. This has the effect of balancing the up-regulated and down-regulated parts of the computation improving the final performance. This has the same effect as imposing adaptively different cutoff thresholds to the up and down regulated genes respectively.

### 5.4 Patients Disease specific active modules

Starting from the filtered list of patient DEG we use the hypergeometric tests to select, with threshold *p <* 0.001, among the co-expression graph modules discovered by Core&Peel the modules that are significantly enriched in up-regulated and down-regulated genes. These two lists of modules constitute the patient-specific active modules.

### 5.5 Drug perturbation enrichment

Each patient-specific active module is used to query the GEO drug perturbation database with the *enrichR* tool, thus obtaining for each module a list of enriched drugs (along with some quantitative measurements of the enrichment). Such lists are then re-organized and indexed using drugs as the main index. In this phase we mark if a drug is enriched for up-regulated or down-regulated genes for the drug perturbations.

### 5.6 Eight scoring functions

For each drug and for the associated significant modules we compute the average value^1^ of the following scoring functions. In all cases higher value correspond to a better countering match of the drug action vs the disease action.

- ’-log10’: is the negative logarithm on base 10 of the p-value returned by enrichR for the module-drug association (used in [Lucchetta and Pellegrini, 2021]).
- ’c score’: is the ‘Combined score’ value returned by enrichR for the module-drug association, that is a metric combining the p-value (from the enrichment test) with a measure of magnitude of enrichment.
- ’deltadeg’, is the difference in the number of genes in the module that come from DEG lists for the patient, taking adversarial pairs (up-vs-down and down-vs-up) as positive, and concordant pairs as negative (used in [Lucchetta and Pellegrini, 2021]).
- ’pval x deltadeg’: is the product of the scores ‘deltadeg’, and ‘-log10’
- ’pval x drug hits’: is the product of the score ‘-log10’ multiplied by the number of drug DEG genes in the module.
- ’pval x deltadeg x drug hits’: is the product of ‘deltadeg’ and ‘pval x drug hits’
- ’odds ratio log’: is the logarithm base 10 of the odds ratio measurement reported by enrichR for the module-drug association
- ’c score x deltadeg’: is the product of ‘c score’ and ‘deltadeg’,

The application of these eight scoring schemes result in eight possible drug rankings for each patient, on which we select the ranking with the highest RHR score.

### 5.7 Method overview for prediction of therapy response

Similarly to the first pipeline (time to event) we start from a list of modules in the modular personal active subnetwork for a given patient. We apply the enrichR package in order to list for each such module the drugs in the GEO database that induce gene expression perturbations significantly enriched in the modules. Also we apply the enrichR package to each module to list the biological processes (either form Gene Ontology - Biological Process, or from the Reactome database) that are significantly enriched in each module. In order the detect the biological processes that are significantly altered in the specific patient we apply the 8 scoring functions defined above so to compute eight rankings of biological processes in a Patient-Biological-Process mapping. This is used as reference background for the subsequent computation that involves both the drugs and the Biological-Process. For each pair of a drug and a biological process we list the modules that are significant for both, and we compute the inner product of the score vectors indexed by the modules in such list. This operation produces a Patient-Drug-Biological-Process mapping. In the final step we compute for each patient-drug pair six derived indices that capture the relative impact of the drug on the top 20 biological processes relevant for that given patient. A schematic figure of the Personalized-Drugrank pipeline for the response derived indices calculations is in Figure 2. The six derived indices are as follows:

- num hits: this index counts the number of top 20 biological processes for the patient that are also in the top 20 biological processes that are relevant for the drug in the patient.
- rhr: this index reports the Reciprocal Hit-Rank (RHR) measure of the top 20 biological process relevant for the patient that are present in the list of the top 20 biological processes relevant for the drug in the patient
- total modules: this index reports the total number of modules that contribute to the num hits index.
- total score: this index reports the sum of the scores for the modules that contribute to the num hits index.
- l2 norm : this index is the L2 norm of vector that has a component for each one of the top 20 biological processes relevant for the patient that are also relevant for the drug on this patient. The value of the vector component is the ratio of the number of modules relevant for the drug and the biological process in the patient over the number of modules relevant for the biological process in the patient.
- l1 norm: this index is the L1 norm of the vector defined above.

**Figure 2:**
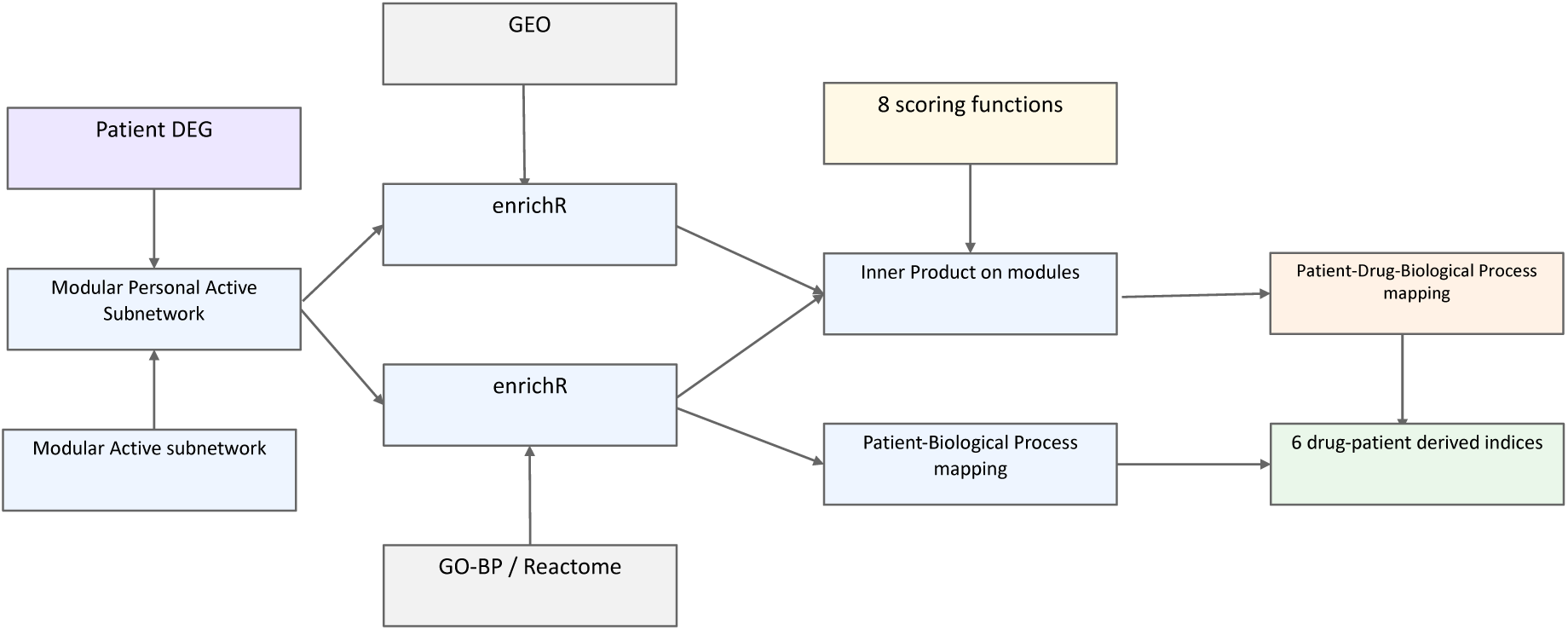
Pipeline for Personalized-DrugRank Index Computations for Response Prediction.

Finally when evaluating the threshold models for response prediction we compute the AUC and maximal accuracy for each derived index and each scoring function and we report the performance of the pair of index and score function with the best AUC. The reference drug reported is one of the drugs that have been administered to the patient and it is present in the GEO database. Tests with KEGG (Kyoto Encyclopedia of Genes and Genomes) and GO-MF (Gene Ontology - Molecular Function) databases were also conducted, which however did result in inferior performance (data not reported).

### 5.8 Cohort Selection

Patient data was retrieved from the National Institute of Health (NIH) National Cancer Institute Genomic Data Commons (GDC) Data Portal using the Cohort Builder tool. The selected patients matched the following criteria:

- *General* :

**–** Program: TCGA
**–** Project: TCGA-COAD, TCGA-READ, TCGA-STAD or TCGA-BRCA projects for colorectal, rectal (from now abbreviated as CRC), stomach (GC) and breast cancer (BC), respectively.
**–** Disease type: for TCGA-BRCA, we selected ductal and lobular neoplasms.
- *Demographic*: alive and dead were both included.
- *Disease Status and History*:

**–** Prior malignancy: no.
**–** Prior treatment: no.
**–** Synchronous malignancy: no.
- *Treatment* :

**–** Treatment type: all treatments were included except surgery and radiation therapy, namely *chemotherapy*, *immunotherapy*, *pharmaceutical* and *targeted molecular therapies*.
**–** Therapeutic agent: therapeutic agent choice was based on most used and its presence in drug perturbation databases. We thus focused our attention on *cyclophosphamide*, *docetaxel*, *paclitaxel*, *doxorubicin hydrochloride* (for BC only), *irinotecan* (for CRC only) and *fluorouracil* (for BC, GC, and CRC).
**–** Treatment outcome: all except *treatment ongoing*, namely *complete response*, *partial response*, *progressive disease*, and *stable disease*.
- *Biospecimen*:

**–** Tissue type: tumor.
**–** Specimen type: solid tissue.
**–** Tumor descriptor: primary.
- *Available Data*:

**–** Data category: transcriptome profiling
**–** Data type: gene expression quantification
**–** Experimental strategy: RNA-Seq
**–** Workflow type: STAR-Counts

The sample sheet and clinical data were then downloaded from the GDC data repository. The resulting general cohorts are defined in Table 4.

**Table 4:**
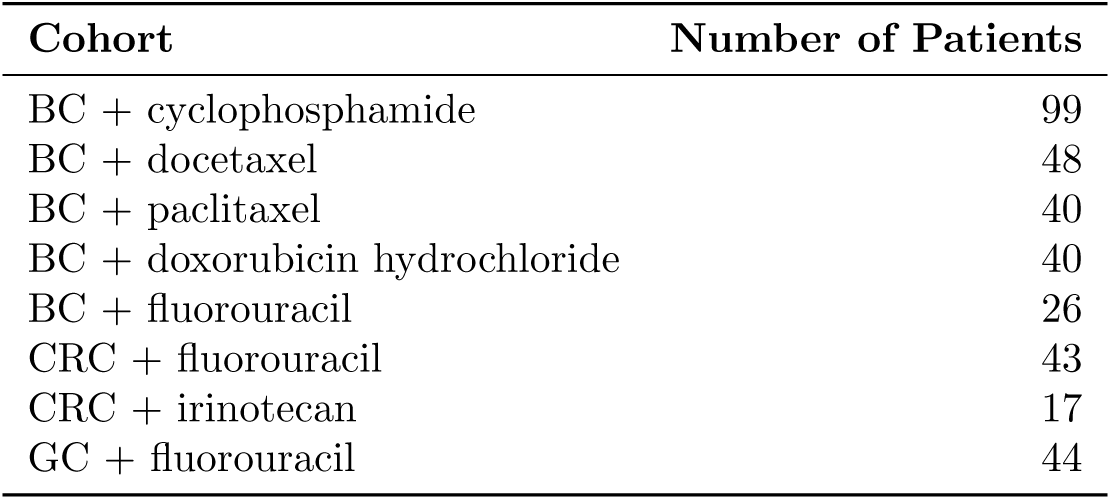
Cohorts. General Cohorts were defined based on cancer type and treatment. BC: breast cancer; CRC: colorectal cancer; GC: gastric cancer (stomach cancer). Clinical features of the cohorts are listed in the Supplementary Materials. Patients often receive more than one drug, thus a patient may belong to more than one cohort.

### 5.9 Identification of Differentially Expressed Genes

#### 5.9.1 Population-level differential expression analysis

Population-level differential expression analysis (DEA) was performed on BC, GC, and CRC RNA-seq data from The Cancer Genome Atlas (TCGA) using the R packages `TCGAbiolinks` and `edgeR sript_population_DEGs`. Gene expression quantification data (workflow type: “STAR - Counts”) were retrieved for both primary tumor and solid tissue normal samples. Raw counts were filtered to retain only relevant sample types, and Ensembl gene identifiers were cleaned to remove version numbers. Normalization was performed using the trimmed mean of M-values (TMM) method, and dispersion estimates were computed within a generalized linear model framework. Differential expression was tested with the quasi-likelihood F-test (glmQLFTest) implemented in `edgeR`. Genes with a false discovery rate (FDR) *<* 0.01 were considered differentially expressed. Ensembl gene IDs were then mapped to HGNC gene symbols using the `biomaRt` package, and lists of significantly upregulated and downregulated genes were generated for subsequent analyses.

#### 5.9.2 Patient-level DEGs

Patient-level DEA was performed for each cohort using a custom R workflow (`script_patient_DEGs.R`). Since matched normal tissue samples were not available for all the patients, we constructed cancer type-specific reference profiles. STAR-aligned count data for “Solid Tissue Normal” samples (*n* = 113 for BC, *n* = 41 for CRC, and *n* = 36 for GC) were retrieved from TCGA selecting the “unstranded” assay for consistency. Expression counts were normalized with `DESeq2` to account for library size differences, and a reference expression profile was generated by computing the mean count per gene across all normal samples.

For each cohort, patient tumor sample identifiers was provided in a CSV file, and analyses were run iteratively. Tumor RNA-seq counts (workflow type: “STAR - Counts”) were retrieved from TCGA using the custom function `query_and_prepare_tumor`, which wraps `TCGAbiolinks’ GDCquery, GDCdownload`, and `GDCprepare` steps. This function includes an automatic retry mechanism with multiple attempts to handle intermittent server errors from the GDC API.

Differential expression was computed with the `perform_DEA` function, which compares counts from a single tumor sample to the reference mean normal profile. If multiple tumor columns were returned for the same patient, only the first was used, and the sample was logged for quality control. For each gene, the log_2_ fold change (logFC) was calculated as:

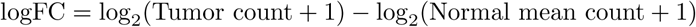

Genes with positive logFC were classified as upregulated, while those with negative logFC were considered downregulated. Ensembl gene identifiers were mapped to HGNC gene symbols using the custom `map_to_hgnc` function, which removes Ensembl version numbers, queries the Ensembl BioMart database, merges annotations with DEA results, and outputs clean, non-redundant gene lists. For each patient, separate lists of upregulated and downregulated genes were exported as text files and stored in a dedicated output directory. Samples processed in previous runs were automatically skipped, and problematic samples (*e.g*., multiple tumor columns) were recorded in a log file.

### 5.10 Personalized-DrugRank Analysis

After reviewing available drug perturbation resources, we selected the Gene Expression Omnibus (GEO) since it includes all the treatments of interest. Active subnetworks for each type of cancer were identified from population-level DEG lists (derived with `edgeR`) using the Core&Peel algorithm [Lucchetta and Pellegrini, 2020].

To extend this framework to patient-level analyses, we developed a batch pipeline based on DrugMerge [Lucchetta and Pellegrini, 2021]. The first phase of the Personalized-DrugRank framework quantifies, for each individual patient, the predicted impact of candidate drugs on the patient-specific disease active subnetwork. This analysis was performed independently for stomach, breast, and colorectal cancer cohorts using cancer-specific scripts with identical computational logic. For each patient, the precomputed lists of upregulated and downregulated genes were used as input. Gene identifiers were harmonized using a curated gene annotation table to ensure compatibility with downstream enrichment and perturbation databases.

#### 5.10.1 Disease-Specific Active Subnetworks

Active subnetworks were previously identified using the Core&Peel community detection algorithm applied to a large gene co-expression network. These subnetworks represent cohesive modules enriched in disease-relevant deregulated genes. Two complementary representations were considered: the largest detected active module (single-module analysis), and the full set of detected modules (multiple-module analysis). These module definitions were cancer-specific but fixed across patients within each cohort.

#### 5.10.2 Enrichment Analysis

Drug-induced transcriptional signatures were retrieved from GEO perturbation libraries using the Enrichr framework. For each patient, enrichment analysis was performed between the patient’s up- and downregulated genes, and drug-induced gene expression signatures. This procedure was carried out separately for the single largest module and for the set of multiple modules. The enrichment step quantifies the statistical association between patient-specific deregulation and drug-induced perturbation profiles, thereby estimating whether a drug is predicted to reverse or reinforce the molecular state of the patient’s tumor.

#### 5.10.3 Construction of Drug Modulation Tables

For each patient and drug database, enrichment results were processed to construct drug modulation tables. These tables summarize:

- the overlap between patient-specific deregulated genes and drug-induced signatures;
- the directionality of modulation (inverse or concordant);
- statistical significance metrics derived from enrichment analysis.

An unfiltered modulation table was first generated for all enriched compounds. Subsequently, p-values were incorporated to produce statistically annotated modulation tables. Filtering procedures were applied to prioritize drugs with statistically significant modulation of the patient-specific active subnetwork. All results were organized hierarchically by cancer type, drug, and patient identifier to maintain reproducibility and traceability.

#### 5.10.4 Computational Execution and Logging

Patients were processed in configurable batches. For each batch, a dedicated log file recorded processing time, analyzed samples, and potential errors, ensuring reproducibility and robustness of execution.

### 5.11 Regression models for survival analysis

The Cox Proportional Hazard analysis [Cox, 1972] is implemented with the *lifelines* python package [Davidson-Pilon, 2024]. For clinical-only features we analyze 14 feature combinations (28 more combinations when subtype is added), for mixed clinical and PDR features we analyze 38 feature combinations (with 90 more combinations when subtype is added). The Cox Proportional Hazard analysis is evaluated with the methods spline (with number of nodes from 2 to 8), piecewise (with two breakpoints) and breslow (semi-parametric). The k-fold cross-validation is done for *k* ranging form 2 to ⌊*n/*2⌋, where *n* is the number of events. For each choice of input features and model parameters, the evaluation is repeated 10 times (or 20 times when the input cohort has fewer than 10 events) with randomization of the fold distributions subject to each fold receiving at least two events (stratified k-fold). Scalar values input parameters are used as scalars. Categorical input parameters are tested both via one-hot encoding and via mapping to an ordered numerical scale, whichever attains the best results. The log-likelihood p-value is measured on a cox model using all the input data. In tables, we report the performance measures for the model attaining the highest mean Concordance Index subject to converging on at least 8 out of 10 (resp. 16 out of 20) repetitions.

### 5.12 Auxiliary Data Sources

TTD Therapeutic Target Database [Zhou et al., 2024].

Clinical guidelines for Colon Cancer [National Comprehensive Cancer Network (NCCN), 2024].

Clinical guidelines for Gastric Cancer (Stomach cancer) [National Comprehensive Cancer Network (NCCN), 2025b]

Clinical guidelines for Breast Cancer [National Comprehensive Cancer Network (NCCN), 2025a].

## Data Availability

All data produced in the present study are available upon reasonable request to the authors

https://github.com/MarcoPellegriniCNR/Personalized-DrugRank

## 6 Acknowledgments

Work funded by the European Union (EU) - Next Generation EU, Mission 4 Component 2 Inv. 1.5 CUP B83C22003930001. Project Tuscany Health Ecosystem (THE).

## Footnotes

1 Here we average on the available cell-line data sets for the same drug.

