## Supplementary Materials for "Joint Prediction of Adjuvant Therapy Response and Time-to-Response for Cancer Patients Using the Personalized-DrugRank Method"

Fiamma Romagnoli* Marco Pellegrini†

March 12, 2026

#### Contents

**A Data and Software availability** 2

**B Tables** 2
B.1 Cohort data 2
B.1.1 BRCA demographic and clinical data 2
B.1.2 CRC demographic and clinical data 4
B.1.3 GC demographic and clinical data 5
B.2 BRCA non-competitive analysis 6
B.2.1 Subpopulation A: Paclitaxel-based 6
B.2.2 Subpopulation B: Cyclophosphamide-based 6
B.2.3 Subpopulation C: Fluorouracil-based 6
B.2.4 Subpopulation D: Docetaxel-based 6
B.2.5 Subpopulation E: Doxorubicin Hydrate-based 6
B.3 STAD non-competitive analysis for Subpopulation Fluorouracil 6
B.4 CRC non-competitive analysis 7
B.4.1 Subpopulation Fluorouracil: model for Complete Remission/Partial Remission 7
B.4.2 Subpopulation Oxaliplatin: model for Complete Remission 7
B.4.3 Subpopulation Oxaliplatin: model for Progressive Disease/Stable Disease 7
B.4.4 Subpopulation Leucovorin: model for Complete Remission 7
B.4.5 Subpopulation Irinotecan: model for Complete Remission/Partial Remission 7
B.4.6 Subpopulation Irinotecan: model for Progressive Disease/Stable Disease 7

**C Role of Cancer Subtypes** 8
C.1 Breast Cancer subtypes 8
C.2 Stomach Cancer subtypes 8
C.3 Colorectal cancer Subtypes 8

**D Choice of Clinical Parameters** 8

**E Relevance of high ranking Biological Processes** 9

*Istituto di Informatica e Telematica del CNR, Via G. Moruzzi 1, 56100-Pisa (Italy).
†Istituto di Informatica e Telematica del CNR, Via G. Moruzzi 1, 56100-Pisa (Italy).

### A Data and Software availability

Data and scripts (both python and R) are posted on Github at
https://github.com/MarcoPellegriniCNR/Personalized-DrugRank.

### B Tables

#### B.1 Cohort data

##### B.1.1 BRCA demographic and clinical data

Table 1: Demographic and clinical data of the BRCA general cohorts

| | | Cyclophosphamide n = 99 | Docetaxel n= 48 | Paclitaxel n = 40 | Doxorubicin n = 40 | Fluorouracil n = 26 |
|---|---|---|---|---|---|---|
| Age at index (years) | Min age | 28 | 30 | 36 | 29 | 28 |
| | Mean age | 53 | 53 | 53 | 50 | 56 |
| | Max age | 78 | 75 | 78 | 78 | 78 |
| Sex, n (%) | Female | 99 (100.0%) | 47 (97.9%) | 39 (97.5%) | 39 (100.0%) | 26 (100.0%) |
| | Male | 0 (0.0%) | 1 (2.1%) | 1 (2.5%) | 0 (0.0%) | 0 (0.0%) |
| Vital status at last follow-up, n (%) | Alive | 91 (91.9%) | 45 (93.8%) | 34 (85.0%) | 33 (84.6%) | 24 (92.3%) |
| | Dead | 8 (8.1%) | 3 (6.2%) | 6 (15.0%) | 6 (15.4%) | 2 (7.7%) |
| Days to last follow-up | Min days | 227 | 216 | 227 | 227 | 300 |
| | Mean days | 1233 | 1165 | 1462 | 1614 | 1474 |
| | Max days | 4159 | 4159 | 4159 | 4159 | 3361 |
| AJCC stage at diagnosis, n (%) | Stage I | 10 (10.1%) | 8 (16.7%) | 3 (7.5%) | 4 (10.3%) | 2 (7.7%) |
| | Stage II | 50 (50.5%) | 23 (47.9%) | 22 (55.0%) | 15 (38.5%) | 15 (57.7%) |
| | Stage III | 31 (31.3%) | 14 (29.2%) | 10 (25.0%) | 15 (38.5%) | 7 (26.9%) |
| | Stage IV | 1 (1.0%) | 0 (0.0%) | 2 (5.0%) | 1 (2.6%) | 0 (0.0%) |
| | Not reported | 7 (7.1%) | 3 (6.2%) | 3 (7.5%) | 4 (10.3%) | 2 (7.7%) |
| Primary diagnosis, n (%) | Infiltrating duct carcinoma, nos | 61 (61.6%) | 35 (72.9%) | 28 (70.0%) | 24 (61.5%) | 13 (50.0%) |
| | Lobular carcinoma, nos | 34 (34.3%) | 12 (25.0%) | 11 (27.5%) | 13 (33.3%) | 11 (42.3%) |
| | Infiltrating duct and lobular carcinoma | 3 (3.0%) | 0 (0.0%) | 0 (0.0%) | 1 (2.6%) | 2 (7.7%) |
| | Infiltrating lobular, mixed | 1 (1.0%) | 0 (0.0%) | 1 (2.5%) | 1 (2.6%) | 0 (0.0%) |
| | Not reported | 0 (0.0%) | 1 (2.1%) | 0 (0.0%) | 0 (0.0%) | 0 (0.0%) |
| | Other | 0 (0.0%) | 0 (0.0%) | 0 (0.0%) | 0 (0.0%) | 0 (0.0%) |

##### B.1.2 CRC demographic and clinical data

Table 2: Demographic and clinical data of the CRC general cohorts

| | | Fluorouracil n = 43 | Irinotecan n = 17 |
|---|---|---|---|
| Age at index (years) | Min age | 42 | 47 |
| | Mean age | 60 | 63 |
| | Max age | 81 | 85 |
| Sex, n (%) | Male | 22 (51.2%) | 13 (76.5%) |
| | Female | 21 (48.8%) | 4 (23.5%) |
| Vital status at last follow-up, n (%) | Alive | 35 (81.4%) | 9 (52.9%) |
| | Dead | 8 (18.6%) | 8 (47.1%) |
| Days to last follow-up | Min days | 0 | 31 |
| | Mean days | 738 | 781 |
| | Max days | 2599 | 1288 |
| AJCC stage at diagnosis, n (%) | Stage I | 2 (4.7%) | 1 (5.9%) |
| | Stage II | 11 (25.6%) | 1 (5.9%) |
| | Stage III | 20 (46.5%) | 3 (17.6%) |
| | Stage IV | 4 (9.3%) | 9 (52.9%) |
| | Not reported | 6 (14.0%) | 3 (17.6%) |
| Primary diagnosis, n (%) | Adenocarcinoma, nos | 28 (65.1%) | 12 (70.6%) |
| | Mucinous adenocarcinoma | 8 (18.6%) | 1 (5.9%) |
| | Papillary adenocarcinoma, nos | 2 (4.7%) | 0 (0.0%) |
| | Carcinoma, nos | 0 (0.0%) | 1 (5.9%) |
| | Not reported | 5 (11.6%) | 3 (17.6%) |

##### B.1.3 GC demographic and clinical data

Table 3: Demographic and clinical data of the GC general cohort

| | | Fluorouracil n = 44 |
|---|---|---|
| Age at index (years) | Min age | 30 |
| | Mean age | 62 |
| | Max age | 77 |
| Sex, n (%) | Male | 33 (75.0%) |
| | Female | 11 (25.0%) |
| Vital status at last follow-up, n (%) | Alive | 24 (54.5%) |
| | Dead | 20 (45.5%) |
| Days to last follow-up | Min days | 1740 |
| | Mean days | 7343 |
| | Max days | 18620 |
| AJCC stage at diagnosis, n (%) | Stage I | 3 (6.8%) |
| | Stage II | 10 (22.7%) |
| | Stage III | 25 (56.8%) |
| | Stage IV | 5 (11.4%) |
| | Not reported | 1 (2.3%) |
| Primary diagnosis, n (%) | Adenocarcinoma, nos | 14 (31.8%) |
| | Adenocarcinoma, intestinal type | 11 (25.0%) |
| | Tubular adenocarcinoma | 11 (25.0%) |
| | Carcinoma, diffuse type | 3 (6.8%) |
| | Mucinous adenocarcinoma | 2 (4.5%) |
| | Other | 3 (6.8%) |

#### B.2 BRCA non-competitive analysis

##### B.2.1 Subpopulation A: Paclitaxel-based

This cohort of breast cancer patients received a therapy based on Cyclophosphamide and Paclitaxel. Breast cancer patients receiving adjuvant cyclophosphamide and paclitaxel typically have early-stage, HER2-negative breast cancer and are treated with a non-anthracycline, taxane-based regimen, often due to intermediate recurrence risk or the need to avoid anthracycline-associated toxicity [National Comprehensive Cancer Network (NCCN), 2025a, page BINV-M 2].

##### B.2.2 Subpopulation B: Cyclophosphamide-based

This cohort of breast cancer patients receiving a therapy based on Cyclophosphamide and other drugs in combination that do not fall into the subpopulations A, C, D and E.

##### B.2.3 Subpopulation C: Fluorouracil-based

This cohort of breast cancer patients received a therapy based on Cyclophosphamide and Fluorouracil. Breast cancer patients receiving adjuvant cyclophosphamide and fluorouracil typically have early-stage, HER2-negative disease, often hormone receptor–positive, and are treated with non-anthracycline CMF-based chemotherapy, frequently due to lower recurrence risk or contraindications to anthracyclines [National Comprehensive Cancer Network (NCCN), 2025a, page BINV-M 2].

##### B.2.4 Subpopulation D: Docetaxel-based

This cohort of breast cancer patients received a therapy based on Cyclophosphamide and Docetaxel. Breast cancer patients receiving adjuvant cyclophosphamide and docetaxel typically have early-stage, HER2-negative HR-negative disease with moderate to high recurrence risk and are treated with a standard, non-anthracycline, taxane-based chemotherapy regimen often chosen to avoid anthracycline toxicity [National Comprehensive Cancer Network (NCCN), 2025a, page BINV-M 3].

##### B.2.5 Subpopulation E: Doxorubicin Hydrate-based

This cohort of breast cancer patients received a therapy based on Cyclophosphamide , Paclitaxel and Doxorubicin Hydrate. Breast cancer patients receiving adjuvant cyclophosphamide, doxorubicin hydrochloride, and paclitaxel generally have early-stage, intermediate- to high-risk disease and are treated with a standard, high-efficacy anthracycline–taxane–alkylating chemotherapy regimen aimed at maximizing recurrence risk reduction while monitoring for cardiotoxicity and neuropathy [National Comprehensive Cancer Network (NCCN), 2025a, page BINV-M 3].

#### B.3 STAD non-competitive analysis for Subpopulation Fluorouracil

This cohort of stomach cancer patients received a therapy based on Fluorouracil and other drugs in combination. Fluoruracil-based therapy is recommended for systemic therapy for unresectable locally advanced, recurrent, or metastatic disease [National Comprehensive Cancer Network (NCCN), 2025b, page GAST-F 4] .

#### B.4 CRC non-competitive analysis

##### B.4.1 Subpopulation Fluorouracil: model for Complete Remission/Partial Remission

Here we consider the subpopulation of TCGA CRC patients receiving only Flurouracil. All these patients achieve Complete Remission/Partial Remission. This cohort of colorectal cancer patients received a therapy based on Fluorouracil that do not fall into the other subpopulations considered. Fluorouracil-based therapies are mainly considered for Stage III colon cancer (node-positive) patients. [National Comprehensive Cancer Network (NCCN), 2024, page COL-D 5]

##### B.4.2 Subpopulation Oxaliplatin: model for Complete Remission

Here we consider the subpopulation of colorectal cancer patients receiving Fluorouracil and Oxaliplatin, but not Irinotecan' or Leucovorin Calcium/Leucovorin. Patients receiving as adjuvant therapy receiving Fluorouracil and Oxaliplatin, but not Irinotecan or Leucovorin Calcium/Leucovorin are typically Stage III (node-positive) patients; or selected high-risk Stage II patients [National Comprehensive Cancer Network (NCCN), 2024, page COL-D 6]

##### B.4.3 Subpopulation Oxaliplatin: model for Progressive Disease/Stable Disease

Here we consider the subpopulation of colorectal cancer patients receiving Fluorouracil and Oxaliplatin, but not Irinotecan' or Leucovorin Calcium/Leucovorin. Patients receiving as adjuvant therapy receiving Fluorouracil and Oxaliplatin, but not Irinotecan or Leucovorin Calcium/Leucovorin are typically Stage III (node-positive) patients; or selected high-risk Stage II patients [National Comprehensive Cancer Network (NCCN), 2024, page COL-D 6]

##### B.4.4 Subpopulation Leucovorin: model for Complete Remission

Here we consider the subpopulation of colorectal cancer patients receiving receiving Fluorouracil and Leucovorin Calcium/Leucovorin, but not Irinotecan. Almost all these patients in this cohort achieve Complete Remission/Partial Remission. This therapy is a post-surgery adjuvant therapy for stage III (and some high-risk stage II) Colorectal cancer patients. [National Comprehensive Cancer Network (NCCN), 2024, page COL-D 5,6]

##### B.4.5 Subpopulation Irinotecan: model for Complete Remission/Partial Remission

This cohort of colorectal cancer patients received a therapy based on Irinotecan in combination with other dugs such as Oxaliplatin, Leucovorin or Fluorouracil. Inrinotecan-based therapies are recommended for systemic therapy for advanced or metastatic disease [National Comprehensive Cancer Network (NCCN), 2024, page COL-D 7]

##### B.4.6 Subpopulation Irinotecan: model for Progressive Disease/Stable Disease

This cohort of stomach cancer patients received a therapy based on Irinotecan in combination with other dugs such as Oxaliplatin, Leucovorin or Fluorouracil. Inrinotecan-based therapies are recommended for systemic therapy for advanced or metastatic disease [National Comprehensive Cancer Network (NCCN), 2024, page COL-D 7]

### C Role of Cancer Subtypes

#### C.1 Breast Cancer subtypes

For Breast Adenocarcinoma we consider the intrinsic/PAM50-like subtypes classification: Her2 (HER2-enriched), LumA (Luminal A) , LumB (Luminal B) , Basal (Basal-like), Normal (Normal-like) [Perou et al., 2000] [Sørlie et al., 2001]. Other subtyping schemes exist that have a rough correspondence to the intrinsic/PAM50-like subtypes classification [Goldhirsch et al., 2013].

We adopted in the Cox regression medeling a ranking of the subtypes in order from the most likely to respond to chemotherapy (paclitaxel) based therapy to the least likely as follows: Basal-like, HER2-enriched, Luminal B, Normal-like, Luminal A [Lehmann et al., 2016] [Schettini et al., 2020] [Martin et al., 2013]. Also we perform Cox regression medeling using one-hot encoding that does not imply any predefined ranking of the subtypes.

#### C.2 Stomach Cancer subtypes

For Stomach (Gastric) cancer we consider the following subtypes classification CIN (Chromosomal Instability), GS (Genomically Stable), MSI (Microsatellite Instability), and EBV(Epstein–Barr Virus) proposed by the TCGA Consortium [Network et al., 2014].

While other classifications have been proposed [Wang et al., 2019] at the moment this classification is still considered the most comprehensive molecular characterization, due to the high number of subject studied (295 primary gastric adenocarcinomas) and the variety of molecular assays used including array-based somatic copy number analysis, whole-exome sequencing, array-based DNA methylation profiling, messenger ribonucleic acid (RNA) sequencing, microRNA (miRNA) sequencing and reverse-phase protein array (RPPA).

Since in our therapy-homogeneous STAD cohort we have a fluorouracil-based therapy we adopted in the Cox regression modelling a ranking of the subtypes in order from the most likely to respond to chemotherapy to the least likely as follows: CIN, MSI, EBV and GS, based on survey of several studies reported in the literature[Sohn et al., 2017]. Also we perform Cox regression modelling using one-hot encoding that does not imply any predefined ranking of the subtypes.

#### C.3 Colorectal cancer Subtypes

For Colorectal Cancer we consider the following subtype classification involved in CRC origin and progression: Chromosomal Instability (CIN); Microsatellite Instability (MSI); and CpG Island Methylator Phenotype (CIMP) [Jass, 2007]. Since the CIMP subtype is not represented in our TCGA cohorts it is not considered in our analysis. Other subtypes classifications exist [Guinney et al., 2015], which however exhibit marked mutual interconnectivity. In our therapy-homogeneous CRC cohort we adopted in the Cox regression medeling a ranking of the subtypes in order from the most likely to respond to therapy to the least likely as follows: CIN, MSI, CIMP [Webber et al., 2015], [Singh et al., 2021]. Also we perform Cox regression medeling using one-hot encoding that does not imply any predefined ranking of the subtypes. Since in our therapy-homogeneous cohort there is a strong prevalence of the CIN subtype (or of no subtype declared) the role of subtypes in therapy response prediction for CRC could not be fully assessed.

### D Choice of Clinical Parameters

The standard clinical parameters included in our study on cancer data correspond to the TNM staging and tumor grade classification (or equivalent) :

- American_Joint_Committee_on_Cancer_Metastasis_Stage
- Neoplasm_Disease_Lymph_Node_Stage_American_Joint_Committee_on_Cancer
- Neoplasm_Disease_Stage_American_Joint_Committee_on_Cancer
- American_Joint_Committee_on_Cancer_Tumor_Stage

In Cox regression models, on all parameters we adopt a standard ranking of increased tumor aggressiveness. For the metastatic parameter we adopt also one-hot encoding in alternative. These four parameters have been chosen since they are the main clinical parameters used in therapy choice and moreover they are common across many tumor types [National Comprehensive Cancer Network (NCCN), 2025a, page ST-1], [National Comprehensive Cancer Network (NCCN), 2025b, page ST-1] and [National Comprehensive Cancer Network (NCCN), 2024, page ST-1]. Other general demographic parameters like patient sex, age and patient ethnicity did not perform well in early experiments in our cohorts and were dropped from further consideration. They should be however be reconsidered when further tumor types are analyzed for which there is evidence of an impact on drug response.

A complementary approach to the one we adopted consists in adding tumor-specific indices (e.g. Gleason scores for Prostate Cancer) for specific tumor types. We leave this line as future research in the refinement of the proposed Personalised-DrugRank method. The example of tumor subtypes as additional feature in the models however indicates that the Personalised-DrugRank is likely to benefit form additional independent relevant signals included in the input features to the prediction models.

### E Relevance of high ranking Biological Processes

One of the key steps in the method for therapy response prediction is to provide a quantitative characterization of the patient cancer in terms of representative biological processes by mapping the patient specific disease active modules onto Gene Ontology or Reactome pathways. As a sanity check we have made an investigation on the terms emerging as most dysregulated for a specific patient and we searched for supporting evidence in the literature for the presence of such terms in high ranking positions. Here we report the ranking an analysis for one such patient. For stomach cancer patient TCGA-3M-AB46 (Male, 70 years old) we have the following TNM/Stage classification:

- American Joint Committee on Cancer Tumor Stage Code: T2B
- American Joint Committee on Cancer Metastasis Stage Code: MX
- Neoplasm Disease Stage American Joint Committee on Cancer Code: STAGE IB
- Neoplasm Disease Lymph Node Stage American Joint Committee on Cancer Code: N0

and cancer subtype CIN-STAD. Thus we expect that chromosomal instability-related pathways are prominent in the pathway ranking obtained from transcriptomic data. In Table 4 we report the list of top ranking biological processes obtained with the score deltadeg, which indeed reports mostly such pathways.
The 30 pathways can be grouped into a few families: Mitotic Division, Chromatid Segregation, Spindle Assembly (GO terms in rank 0–8, 11–14, 16–22, 27–28, 30). DNA Metabolism, DNA Replication, DNA Repair, DNA Damage Response (GO terms in ranks 1, 9–10, 15, 23–26, 29). Cell Cycle Progression, Checkpoints (GO terms in ranks 5, 16–19, 24, 26, 30). These pathways are known to be dysregulated in gastric cancer (in particular for the CIN subtype). See e.g. [Network et al., 2014], [Nemtsova et al., 2023], [Grabsch et al., 2003], [Kamran et al., 2017], [Chen et al., 2022], [Sahgal et al., 2023], [da Costa et al., 2023], [Guan et al., 2021], [Chen et al., 2018].

Table 4: Ranking of Gene Ontology terms in dysregulated active cancer modules for patient TCGA-3M-AB46

| rank | GO pathway | deltadeg | n. modules |
|---|---|---|---|
| 0 | Mitotic Sister Chromatid Segregation GO:0000070 | 6236.0 | 205 |
| 1 | DNA Metabolic Process GO:0006259 | 5751.0 | 189 |
| 2 | Sister Chromatid Segregation GO:0000819 | 5474.0 | 159 |
| 3 | Mitotic Spindle Organization GO:0007052 | 5370.0 | 151 |
| 4 | Mitotic Nuclear Division GO:0140014 | 5332.0 | 152 |
| 5 | Positive Regulation of Cell Cycle Process GO:0090068 | 5298.0 | 144 |
| 6 | Mitotic Metaphase Chromosome Alignment GO:0007080 | 5287.0 | 141 |
| 7 | Metaphase Chromosome Alignment GO:0051310 | 5192.0 | 138 |
| 8 | Microtubule Cytoskeleton Organization Involved in Mitosis GO:1902850 | 4961.0 | 127 |
| 9 | DNA Repair GO:0006281 | 4943.0 | 136 |
| 10 | DNA Damage Response GO:0006974 | 4827.0 | 143 |
| 11 | Mitotic Spindle Assembly GO:0090307 | 4822.0 | 124 |
| 12 | Mitotic Spindle Elongation GO:0000022 | 4530.0 | 113 |
| 13 | Mitotic Spindle Midzone Assembly GO:0051256 | 4530.0 | 113 |
| 14 | Spindle Assembly GO:0051225 | 4185.0 | 99 |
| 15 | Replication Fork Processing GO:0031297 | 4147.0 | 106 |
| 16 | Negative Regulation of Mitotic Metaphase/Anaphase Transition GO:0045841 | 4134.0 | 107 |
| 17 | Mitotic Spindle Checkpoint Signaling GO:0071174 | 4072.0 | 104 |
| 18 | Spindle Assembly Checkpoint Signaling GO:0071173 | 4072.0 | 104 |
| 19 | Mitotic Spindle Assembly Checkpoint Signaling GO:0007094 | 4072.0 | 104 |
| 20 | Mitotic Cytokinesis GO:0000281 | 3896.0 | 97 |
| 21 | Positive Regulation of Cytokinesis GO:0032467 | 3817.0 | 91 |
| 22 | Cytoskeleton-Dependent Cytokinesis GO:0061640 | 3792.0 | 93 |
| 23 | DNA-templated DNA Replication GO:0006261 | 3755.0 | 94 |
| 24 | Regulation of Cell Cycle Checkpoint GO:1901976 | 3525.0 | 87 |
| 25 | DNA Recombination GO:0006310 | 3497.0 | 83 |
| 26 | Regulation of DNA Damage Checkpoint GO:2000001 | 3412.0 | 82 |
| 27 | Centromere Complex Assembly GO:0034508 | 3357.0 | 79 |
| 28 | Establishment of Chromosome Localization GO:0051303 | 3346.0 | 86 |
| 29 | DNA Replication GO:0006260 | 3235.0 | 109 |
| 30 | Regulation of Mitotic Cell Cycle Phase Transition GO:1901990 | 3232.0 | 70 |
